# Treatment of Slow-flow After Primary Percutaneous Coronary Intervention With Flow-mediated Hyperemia. The Randomized RAIN-FLOW Study

**DOI:** 10.1101/2023.02.21.23286266

**Authors:** Josep Gomez-Lara, Montserrat Gracida, Fernando Rivero, Alejandro Gutiérrez, Guillem Muntané-Carol, Rafael Romaguera, Lara Fuentes, Ana Marcano, Gerard Roura, José Luis Ferreiro, Luis Teruel, Salvatore Brugaletta, Fernando Alfonso, Josep Comín-Colet, Joan-Antoni Gomez-Hospital

## Abstract

**Background:** ST-segment Elevation Myocardial Infarction (STEMI) complicated with no reflow after primary percutaneous coronary intervention is associated with adverse outcomes. Although several hyperemic drugs have shown to improve the Thrombolysis In Myocardial Infarction (TIMI) flow, optimal treatment of no reflow remains unsettled. Saline infusion at 20 ml/min via a dedicated microcatheter causes (flow-mediated) hyperemia. The objective is to compare the efficacy of pharmacologic *vs*. flow-mediated hyperemia in STEMI patients complicated with no reflow.

**Methods:** STEMI patients with no reflow were randomized to receive either adenosine or nitroprusside *vs*. flow-mediated hyperemia. The angiographic corrected TIMI Frame Count (cTFC) and the Minimal Microcirculatory Resistance (MMR), as assessed with intracoronary pressure-thermistor wire, dedicated microcatheter and thermodilution techniques, were compared after study interventions.

**Results:** Sixty-seven were included (30 allocated to pharmacologic and 37 to flow-mediated hyperemia). After study interventions, cTFC (40.2±23.1 *vs*. 39.2±20.7; p=0.858) and MMR (753.6±661.5 *vs*. 993.3±740.8 Wood units; p=0.174) were similar between groups. TIMI 3 flow was observed in 26.7% *vs*. 27.0% (p=0.899). Flow-mediated hyperemia showed two different thermodilution patterns during saline infusion indicative of the severity of the no reflow phenomenon. In-hospital death and non-fatal heart failure were observed in 10.4% and 26.9%, respectively.

**Conclusions:** Both treatments showed similar (and limited) efficacy restoring coronary flow. Flow-mediated hyperemia with thermodilution pattern assessment allowed the simultaneous characterization of the no reflow degree and response to hyperemia. No reflow was associated with a high rate of adverse outcomes. Further research is warranted to prevent and to treat no reflow in STEMI patients (NCT04685941).

## Introduction

Current treatment of ST-segment Elevation Myocardial Infarction (STEMI), including emergent Primary Percutaneous Coronary Intervention (PPCI), has drastically improved the survival of patients admitted for STEMI ^1, 2^. However, despite best care interventions, in-hospital mortality remains around 5% ^1, 2^. Cardiogenic shock is the main cause of death in STEMI patients (>50%) ^3, 4^. Predictors of cardiogenic shock are patient’s age, diabetes mellitus, heart failure, culprit lesions located in large vessels and the presence of no reflow (or slow flow) after stent implantation ^3^.

The no reflow phenomenon is defined as the absence of normal flow (compared to the other coronary arteries) after appropriate culprit (epicardial) vessel revascularization ^5^. It is indicative of an unsuccessful microcirculatory reperfusion ^5^. Usually, no reflow is classified according to the Thrombolysis In Myocardial Infarction (TIMI) flow between 0 and 2; and is observed in around 10-30% of STEMI patients undergoing PPCI ^6^. Although multiple pharmacological and mechanical interventions have been investigated to prevent and treat the no reflow phenomenon, none of those interventions have been shown effective to reduce the infarcted area ^5, 7^. Intracoronary infusion of microcirculatory vasodilators (such as nitroprusside and adenosine) is often used to treat no reflow in the clinical practice ^5^. However, according to the current practice guidelines, bail-out administration of IIb/IIIa glycoprotein inhibitors is the only recommended strategy when no reflow is observed after PPCI ^8, 9^.

In patients with chronic coronary syndromes, intracoronary infusion of saline at 15-30 ml/min via a dedicated microcatheter has been demonstrated to cause (flow-mediated) hyperemia in a similar or superior degree than intracoronary or intravenous adenosine administration ^10^. Flow-mediated hyperemia is often achieved at 15 seconds of the beginning of saline infusion ^10^. The safety and efficacy of flow-mediated hyperemia to treat the no reflow phenomenon in STEMI patients is unknown. The objective of the present study is to investigate the safety and efficacy of flow-mediated hyperemia, as compared with standard-of-care pharmacologic-mediated hyperemia, in STEMI patients presenting with slow flow after PPCI.

## Methods

### Endpoints

The present study has two co-primary endpoints: to compare the corrected TIMI Frame Count (cTFC) and the thermodilution-based Minimal Microcirculatory Resistance (MMR) after treatment of no reflow between the two study groups. A detailed description of the co-primary endpoints and secondary endpoints is shown in the **Supplementary Appendix**. All study endpoints have been assessed by a central core-laboratory (Barcelona Cardiac Imaging core-laboratory; BARCICORE-lab, Spain) blinded to the study allocation.

### Study design and population

The RAIN-FLOW study (NCT 04685941) is an investigator-initiated, proof-of-concept, two-arms, randomized, and multi-center study. The study was performed according to the provisions of the Declaration of Helsinki; and the ethics committee of each participating center approved the study protocol. The ethics committee of the University Hospital of Bellvitge acted as referring ethics committee. Oral consent was mandated (as per protocol), and all participants signed written informed consent after the procedure. All authors had access to the study data and the corresponding author takes responsibility for its integrity and the data analysis.

STEMI patients undergoing PPCI within 12 hours of symptom’s onset and presenting with sustained slow coronary flow after stent implantation (or stent post-dilatation) were eligible for the study. Patients with cardiogenic shock, high bleeding risk (including previous stroke, creatinine clearance < 30 ml/min and active bleeding), visualization of distal thrombus embolization with several occluded branches of the study vessel, stent thrombosis or culprit lesion located in coronary bypass were not eligible. Patients accepting to participate were 1:1 randomized to one of the study interventions for slow flow treatment: 1) standard of care pharmacologic-mediated hyperemia with intracoronary adenosine or nitroprusside or 2) treatment with flow-mediated hyperemia with saline infusion via a dedicated microcatheter located in the proximal segment of the culprit vessel. Randomization was performed electronically using computer generated random algorithms.

The sample size was estimated to demonstrate superiority of the experimental arm for both co-primary endpoints. According to the sample size calculation, a total of 100 patients were needed (50 per group). The **Supplementary Appendix** reports the assumptions of the sample size calculation. However, after nearly 2 years of inclusion, the study showed a slow recruitment rate and only 2/3 of the pre-specified population had been recruited. For this reason, the Steering Committee decided to perform an unplanned interim analysis of the study results. According to the interim analysis, the study hypotheses of both co-primary endpoints were unlikely to be achieved with the estimated sample size. Moreover, a re-calculation of the sample size using a non-inferiority study design (using the data of the present study) showed that > 100 patients per group were needed. Considering the slow recruitment rate and the results of the interim analysis, it was decided to terminate the study prematurely.

### Study interventions

Study interventions are detailed in the **Supplementary appendix** and are summarized in **Figure 1**. As per protocol, all patients were randomized to one of the two study interventions: Pharmacologic-mediated hyperemia or Flow-mediated hyperemia. Flow-mediated hyperemia was performed with intracoronary saline infusion via a dedicated micro-catheter (Ray Flow, Hexacath, France) at 20 ml/min for 135 seconds. Continuous recording of the absolute coronary blood flow (ACBF) and MMR was assessed using a dedicated pressure-thermistor coronary wire (Pressurewire X, Abbott, United States) and software (Coroflow, Abbott, United States) as appropriate ^11-13^

**Figure 1.**
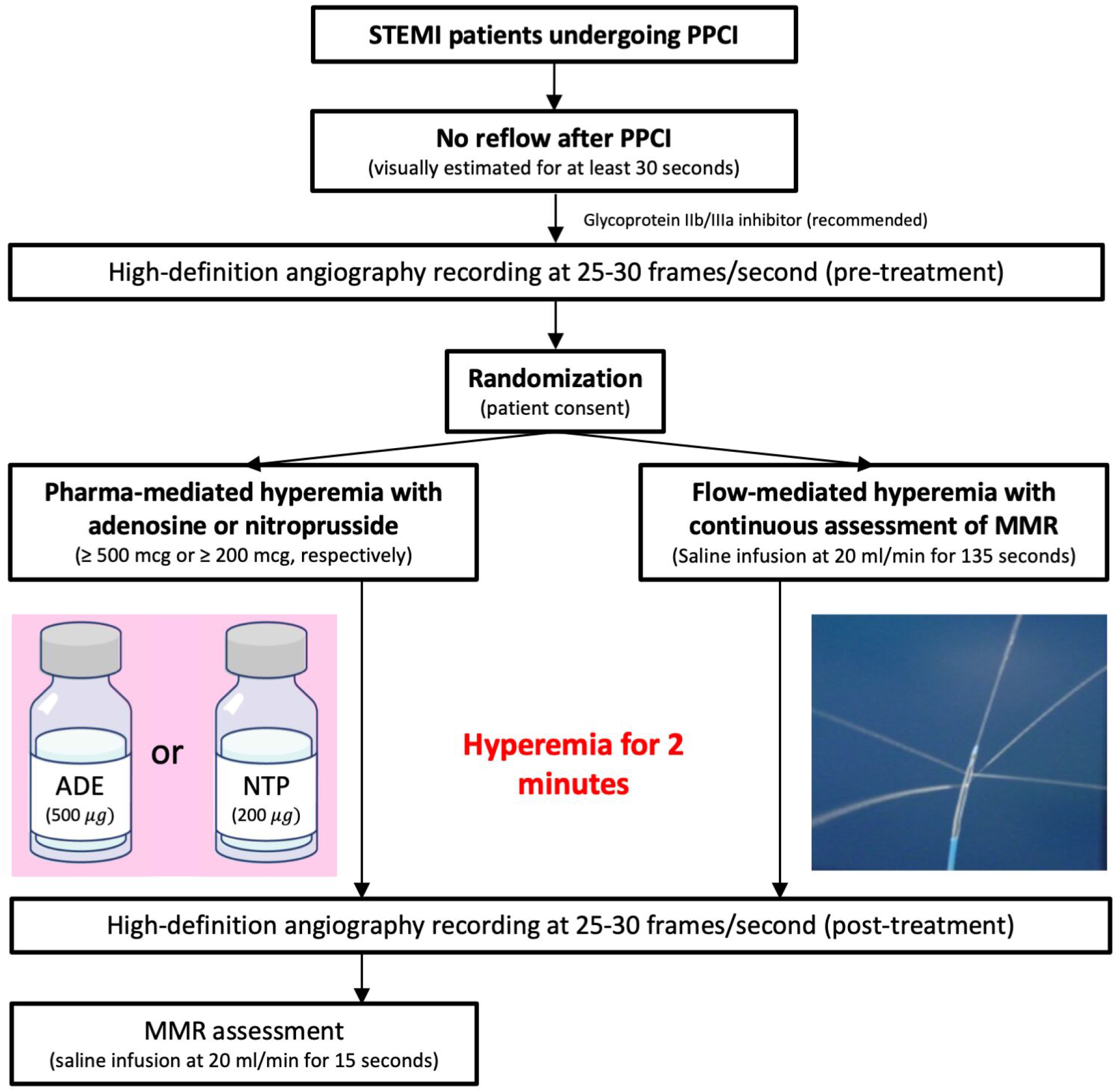
Study interventions (as per protocol). ADE= Adenosine; MMR= Minimal Microcirculatory Resistance; NTP= Nitroprusside; PPCI= Primary Percutaneous Coronary Intervention.

### Statistical analysis

Categorical variables were presented as counts and percentages, and quantitative variables as mean ± standard deviation. Continuous variables were tested for normal distribution with the Kolmogorov-Smirnov test. Comparisons of categorical variables were estimated with the chi-square test and comparisons of quantitative values between groups were estimated with the Student T-test for paired and non-paired samples as appropriate. A two-sided p value <0.05 was considered statistically significant. Statistical analysis was performed with the SPSS software, version 20.0 (SPSS Inc., IL, USA).

## Results

### Patients

From January 23^rd^, 2021, to November 9^th^, 2022, a total of 1257 patients underwent to PPCI in 3 participating Institutions. A total of 132 patients with slow flow fulfilled the inclusion criteria, and 67 were included in the study (30 patients were allocated to pharmacologic and 37 to flow-mediated hyperemia treatment). Most of the eligible patients not included in the study were admitted during non-working hours, especially at nighttime. The flow chart of the study is shown in **Figure 2**.

**Figure 2.**
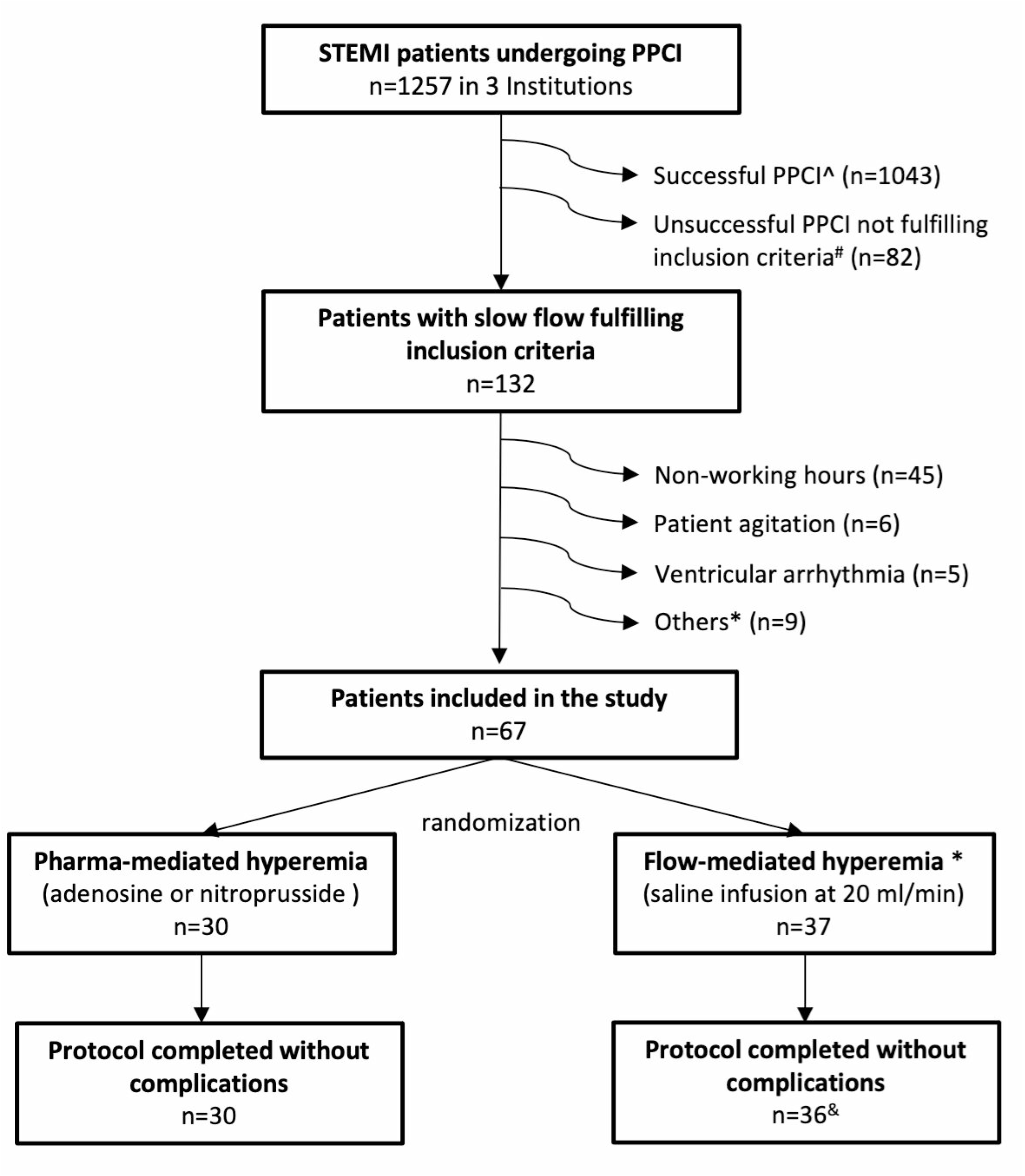
Study flow chart PPCI= Primary Percutaneous Coronary Intervention; TIMI= Thrombolysis In Myocardial Infarction. ^ Successful PPCI was defined as final TIMI 3 flow. # Patients with unsuccessful PPCI not suitable for the study were mostly: subacute myocardial infarction, end-stage renal dysfunction, distal thrombus embolization during PPCI, patients resuscitated from out-of-hospital cardiac arrest, and cardiogenic shock. Those patients were not registered in the screening failure list. * Other causes of screening failure included urgency to end the procedure for laboratory demand (n=4), technical issues with the infusion pump (n=2), patient refuse to participate (n=2) and included in other clinical trial (n=1). & One patient of the flow-mediated hyperemia group presented with coronary dissection during the advance of pressure wire and required a stent implantation. This patient completed the study protocol after treatment of the coronary dissection.

### Clinical, angiographic, and procedural characteristics

Both groups presented with similar baseline characteristics. **Table 1** shows the clinical baseline characteristics of the study groups. The mean age was 67.7±12.3 years and 76.1% were men. Clinical, angiographic, and procedural characteristics of the STEMI treatment are shown in **Table 2**. Both groups presented with similar reperfusion time (median of 250 minutes from symptom’s onset to PCI; interquartile range: 150 to 525 minutes), and STEMI location (53.7% of patients had anterior ST-segment elevation). At hospital admission, heart failure was present in 28.4% of patients with a trend towards lower percentage of patients with Killip class >1 in the pharmacologic than in the flow-mediated hyperemia group (16.6% vs. 37.8%; p=0.155).

**Table 1.** Baseline clinical characteristics

|  | **Pharma-mediated**<br>**hyperemia**<br>**(n=30)** | **Flow-mediated**<br>**hyperemia**<br>**(n=37)** | **p** |
| --- | --- | --- | --- |
| **Age, years** | 65.7±10.5 | 69.4±13.5 | 0.222 |
| **Males** | 25 (83.3%) | 26 (70.3%) | 0.212 |
| **Body mass index** | 28.0±4.4 | 27.5±4.0 | 0.604 |
| **Smoking status:** |  |  | 0.068 |
| Never | 9 (30.0%) | 20 (54.1%) |  |
| Former | 14 (46.7%) | 8 (21.6%) |  |
| Current | 7 (23.3%) | 9 (24.3%) |  |
| **Hypertension** | 16 (53.3%) | 22 (59.5%) | 0.615 |
| **Hypercholesterolemia** | 19 (63.3%) | 21 (56.8%) | 0.585 |
| **Diabetes mellitus** | 11 (36.7%) | 14 (38.9%) | 0.853 |
| Insulin-treated | 3 (10.0%) | 6 (16.2%) | 0.458 |
| **Comorbidities:** |  |  |  |
| CKD | 4 (13.3%) | 11 (29.7%) | 0.109 |
| COPD | 1 (3.3%) | 2 (5.4%) | 0.683 |
| PAD | 0 | 2 (5.4%) | 0.196 |
| Previous PCI | 4 (13.3%) | 1 (2.7%) | 0.100 |
| **Chronic treatment:** |  |  |  |
| Antiplatelet therapy | 2 (6.7%) | 5 (13.5%) | 0.362 |
| B-blocker | 4 (13.3%) | 6 (16.2%) | 0.742 |
| ACEI/ARB | 16 (53.3%) | 18 (48.6%) | 0.703 |
| Statin | 11 (36.7%) | 9 (24.3%) | 0.272 |
| Oral anticoagulation | 2 (6.7%) | 3 (8.1%) | 0.823 |

ACEI= Angiotensin-Converting Enzyme Inhibitors; ARB= Angiotensin II Receptor Blockers; CKD= Chronic Kidney Disease (creatinine clearance <60 ml/min); COPD= Chronic Obstructive Pulmonary Disease; MI= myocardial infarction; PAD= Peripheral Artery Disease; PCI= Percutaneous Coronary Intervention

**Table 2.**
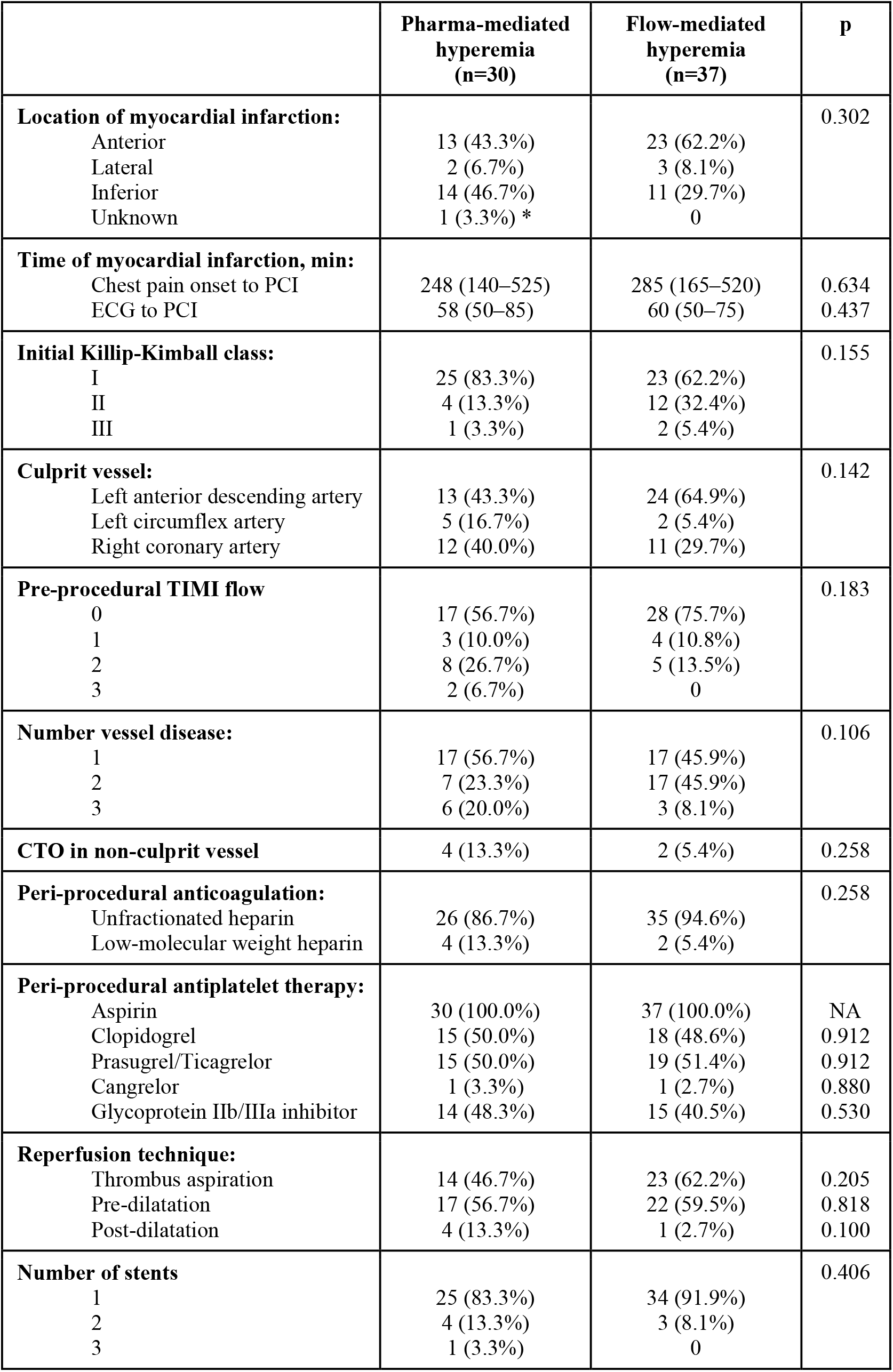
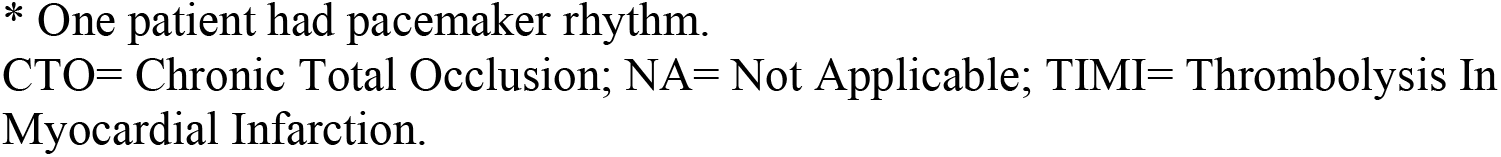
Clinical, angiographic, and procedural characteristics of the myocardial infarction treatment.

|  | Pharma-mediated<br>hyperemia<br>(n=30) | Flow-mediated<br>hyperemia<br>(n=37) | p |
| --- | --- | --- | --- |
| **Location of myocardial infarction:** |  |  | 0.302 |
| Anterior | 13 (43.3%) | 23 (62.2%) |  |
| Lateral | 2 (6.7%) | 3 (8.1%) |  |
| Inferior | 14 (46.7%) | 11 (29.7%) |  |
| Unknown | 1 (3.3%) * | 0 |  |
| **Time of myocardial infarction, min:** |  |  |  |
| Chest pain onset to PCI | 248 (140–525) | 285 (165–520) | 0.634 |
| ECG to PCI | 58 (50–85) | 60 (50–75) | 0.437 |
| **Initial Killip-Kimball class:** |  |  | 0.155 |
| I | 25 (83.3%) | 23 (62.2%) |  |
| II | 4 (13.3%) | 12 (32.4%) |  |
| III | 1 (3.3%) | 2 (5.4%) |  |
| **Culprit vessel:** |  |  | 0.142 |
| Left anterior descending artery | 13 (43.3%) | 24 (64.9%) |  |
| Left circumflex artery | 5 (16.7%) | 2 (5.4%) |  |
| Right coronary artery | 12 (40.0%) | 11 (29.7%) |  |
| **Pre-procedural TIMI flow** |  |  | 0.183 |
| 0 | 17 (56.7%) | 28 (75.7%) |  |
| 1 | 3 (10.0%) | 4 (10.8%) |  |
| 2 | 8 (26.7%) | 5 (13.5%) |  |
| 3 | 2 (6.7%) | 0 |  |
| **Number vessel disease:** |  |  | 0.106 |
| 1 | 17 (56.7%) | 17 (45.9%) |  |
| 2 | 7 (23.3%) | 17 (45.9%) |  |
| 3 | 6 (20.0%) | 3 (8.1%) |  |
| **CTO in non-culprit vessel** | 4 (13.3%) | 2 (5.4%) | 0.258 |
| **Peri-procedural anticoagulation:** |  |  | 0.258 |
| Unfractionated heparin | 26 (86.7%) | 35 (94.6%) |  |
| Low-molecular weight heparin | 4 (13.3%) | 2 (5.4%) |  |
| **Peri-procedural antiplatelet therapy:** |  |  |  |
| Aspirin | 30 (100.0%) | 37 (100.0%) | NA |
| Clopidogrel | 15 (50.0%) | 18 (48.6%) | 0.912 |
| Prasugrel/Ticagrelor | 15 (50.0%) | 19 (51.4%) | 0.912 |
| Cangrelor | 1 (3.3%) | 1 (2.7%) | 0.880 |
| Glycoprotein IIb/IIIa inhibitor | 14 (48.3%) | 15 (40.5%) | 0.530 |
| **Reperfusion technique:** |  |  |  |
| Thrombus aspiration | 14 (46.7%) | 23 (62.2%) | 0.205 |
| Pre-dilatation | 17 (56.7%) | 22 (59.5%) | 0.818 |
| Post-dilatation | 4 (13.3%) | 1 (2.7%) | 0.100 |
| **Number of stents** |  |  | 0.406 |
| 1 | 25 (83.3%) | 34 (91.9%) |  |
| 2 | 4 (13.3%) | 3 (8.1%) |  |
| 3 | 1 (3.3%) | 0 |  |

\* One patient had pacemaker rhythm.
CTO= Chronic Total Occlusion; NA= Not Applicable; TIMI= Thrombolysis In Myocardial Infarction.

TIMI flow at randomization (observed after stent implantation or post-dilatation and before no reflow treatment) was similarly observed in both groups (**Table 3**). TIMI flow 0 was observed in 4.5% of patients (3.3% pharmacologic *vs*. 5.4% flow-mediated group), TIMI flow 1 in 40.3% (43.3% *vs*. 37.8%) and TIMI flow 2 in 55.2% (53.3% *vs*. 56.8%, respectively); p=0.853. The pre-intervention angiographic cTFC was also similar between groups (59.3±26.7 *vs*. 55.1±28.3; p=0.552).

**Table 3.** Angiographic and ECG results before and after slow flow treatment.

|  | Pharma-mediated<br>hyperemia<br>(n=30) | Flow-mediated<br>hyperemia<br>(n=37) | p |
| --- | --- | --- | --- |
| **TIMI flow before slow-flow treatment** |  |  | 0.853 |
| 0 | 1 (3.3%) | 2 (5.4%) |  |
| 1 | 13 (43.3%) | 14 (37.8%) |  |
| 2 | 16 (53.3%) | 21 (56.8%) |  |
| 3 | 0 | 0 |  |
| **TIMI flow after slow-flow treatment** |  |  | 0.899 |
| 0 | 0 | 0 |  |
| 1 | 3 (10.0%) | 5 (13.5%) |  |
| 2 | 19 (63.3%) | 22 (59.5%) |  |
| 3 | 8 (26.7%) | 10 (27.0%) |  |
| **Angiographic cTFC, n:** |  |  |  |
| Before slow-flow treatment* | 59.3±26.7 | 55.1±28.3 | 0.552 |
| After slow-flow treatment | 40.2±23.1 | 39.2±20.7 | 0.858 |
| Delta | 22.4±29.5 | 15.2±18.2 | 0.248 |
| Comparison before–after (p value) | <0.001 | <0.001 | NA |
| **Angiographic myocardial blush (post), n:** |  |  | 0.853 |
| 0 | 11 (36.7%) | 10 (27.0%) |  |
| 1 | 14 (46.7%) | 20 (54.1%) |  |
| 2 | 4 (13.3%) | 6 (16.2%) |  |
| 3 | 1 (3.3%) | 1 (2.7%) |  |
| **ECG maximal ST elevation, mm #** |  |  |  |
| Before PCI | 3.9±2.4 | 3.5±2.4 | 0.474 |
| After PCI | 1.4±0.83 | 1.8±1.1 | 0.117 |
| **ECG maximal ST resolution, % #** |  |  |  |
| ≥50% | 18 (69.2%) | 18 (51.4%) | 0.162 |
| ≥70% | 9 (34.6%) | 7 (20.0%) | 0.199 |

cTFC= corrected TIMI frame count; ECG= Electrocardiogram; TIMI= Thrombolysis In Myocardial Infarction.
\* TIMI frame count was not estimated in 6 patients (2 in the pharma and 4 in the saline-induced hyperemia group) because contrast did not reach the distal landmark for TIMI frame count analysis.

### Slow-flow treatment

All patients included in the study successfully underwent to the allocated intervention. A total of 30 patients underwent to pharmacologic-mediated hyperemia with intracoronary adenosine (n=11; mean dose of 591±170 mcg), nitroprusside (n=11; mean dose of 464±103 mcg) or a combination of both agents (n=8, doses of 405±238 and 356±140 mcg, respectively); and 37 patients underwent to flow-mediated hyperemia with saline infusion. One patient in this group presented with proximal dissection of the left circumflex artery caused by the pressure wire. This patient was treated according to the study protocol (with flow-mediated hyperemia) after treatment of the coronary dissection with stent implantation without further complications.

The mean time required to start the study interventions was statistically significant shorter in the pharmacologic than in the flow-mediated hyperemia group (3.7±3.3 *vs*. 5.9±4.3 minutes; p=0.025). A total of 40% of patients undergoing flow-mediated hyperemia were also treated with hyperemic drugs but only after all study interventions were finalized, as bail-out, due to persistent slow flow.

### End points

The study endpoints observed after slow flow treatment are shown in **Tables 3 and 4** and are summarized in **Figure 3**. The cTFC was improved in both arms between pre and post study interventions. In the pharmacologic-mediated hyperemia group, cTFC was reduced from 59.3±26.7 to 40.2±23.1 frames (p<0.001); and in the flow-mediated hyperemia group from 55.1±28.3 to 39.2±20.7 frames (p<0.001). There were no statistically significant differences regarding the post treatment cTFC (p=0.858) and the delta change cTFC (p=0.248) between groups.

**Table 4.** Physiologic results after slow flow treatment.

|  | Pharma-mediated hyperemia at 15 sc. (n=30) | Flow-mediated hyperemia group (n=37) |  | p<sup>^</sup> | p<sup>*</sup> | p<sup>#</sup> |
| --- | --- | --- | --- | --- | --- | --- |
|  |  | At 15 sc. | At 135 sc. |  |  |  |
| **Pressure at hyperemia, mmHg:**<br>Aortic<br>Distal | 84.0±18.9<br>81.4±19.5 | 82.5±19.6<br>77.0±20.1 | 82.4±18.1<br>77.0±18.7 | 0.901<br>1.000 | 0.747<br>0.372 | 0.715<br>0.353 |
| **Fractional Flow Reserve, value** | 0.97±0.05 | 0.93±0.07 | 0.93±0.08 | 0.254 | 0.021 | 0.048 |
| **Absolute coronary blood flow, ml/min**<br>Normalized value | 161.8±101.1<br>166.2±105.8 | 149.6±122.5<br>159.8±130.4 | 117.4±84.0<br>121.6±82.0 | <0.001<br>0.001 | 0.665<br>0.828 | 0.056<br>0.058 |
| **Minimal microcirculatory resistance, WU** | 753.6±661.5 | 849.9±702.0 | 993.3±740.8 | <0.001 | 0.571 | 0.174 |

WU= Wood Units
<sup>^</sup> p value indicates the paired differences of physiologic parameters at 15 and at 135 seconds in the flow-mediated hyperemia group; <sup>\*</sup> p value indicates the difference between the study groups at 15 seconds of the saline infusion; <sup>#</sup> p value indicates the difference between the physiologic results obtained at 15 seconds in the pharmacologic and at 135 seconds in the flow-mediated hyperemia group.

**Figure 3.**
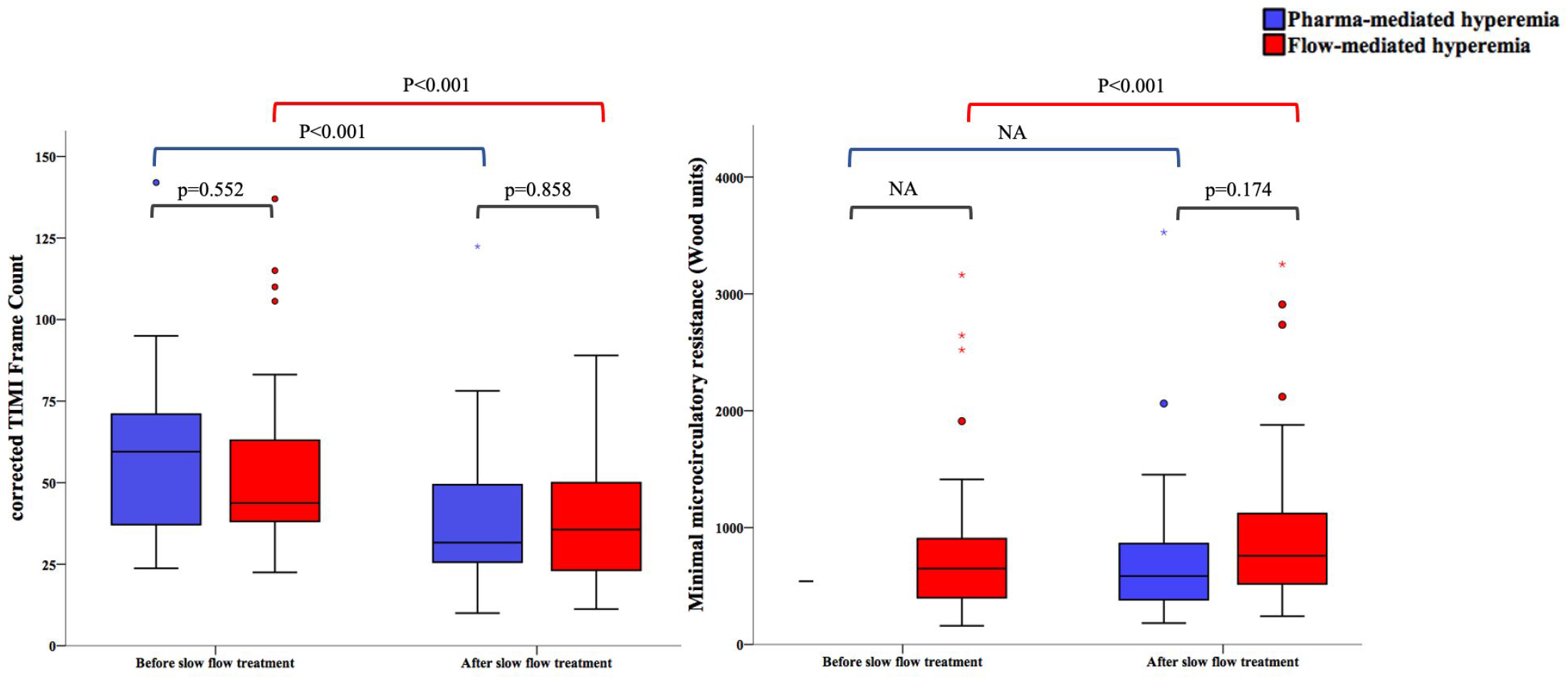
Primary endpoints. Boxplot of the study endpoints before and after treatment. Of note, MMR was not assessed before treatment in the pharmacologic group. In this group, MMR was assessed after administration of hyperemic drugs. In the flow-mediated hyperemia group, MMR values were assessed during saline infusion at 15 (before treatment) and at 135 seconds (after treatment). MMR= Minimal Microcirculatory Resistance; TIMI=Thrombolysis In Myocardial Infarction

MMR after study interventions (obtained at 15 seconds in the pharmacologic and at 135 seconds in the flow-mediated hyperemia group) was numerically lower in the pharmacologic (753.6±661.5 Wood units) than in the flow-mediated hyperemia group (993.3±740.8); p=0.174.

In patients treated with flow-mediated hyperemia, MMR worsened from the beginning (849.9±702.0 Wood units measured at 15 seconds) to the end of saline infusion (993.3±740.8 Wood units measured at 135 seconds); p<0.001. MMR changes were explained by different distal temperature values, indicative of the temperature of the mixed blood and the infused saline at room temperature, observed at the beginning (−0.53º) and at the end (−0.65º) of the saline infusion. However, two different thermodilution patterns were observed during saline infusion in this group. **Figure 4** shows the two different thermodilution patterns observed during saline infusion in the flow-mediated hyperemia group (appropriate *vs*. insufficient saline clearance patterns). Appropriate saline clearance pattern was considered when the distal temperature was maintained steady during the saline infusion. In contrast, insufficient saline clearance pattern was defined by a progressive distal temperature decrease (> 2 standard deviation of the delta value between the distal temperature at beginning and at the end of the saline infusion: -0.44º). In this pattern, the progressive drop of the distal temperature was explained by a deficient clearance of the infused saline (at 20 ml/min) that accumulates in the distal segment of the treated vessel.

**Figure 4.**
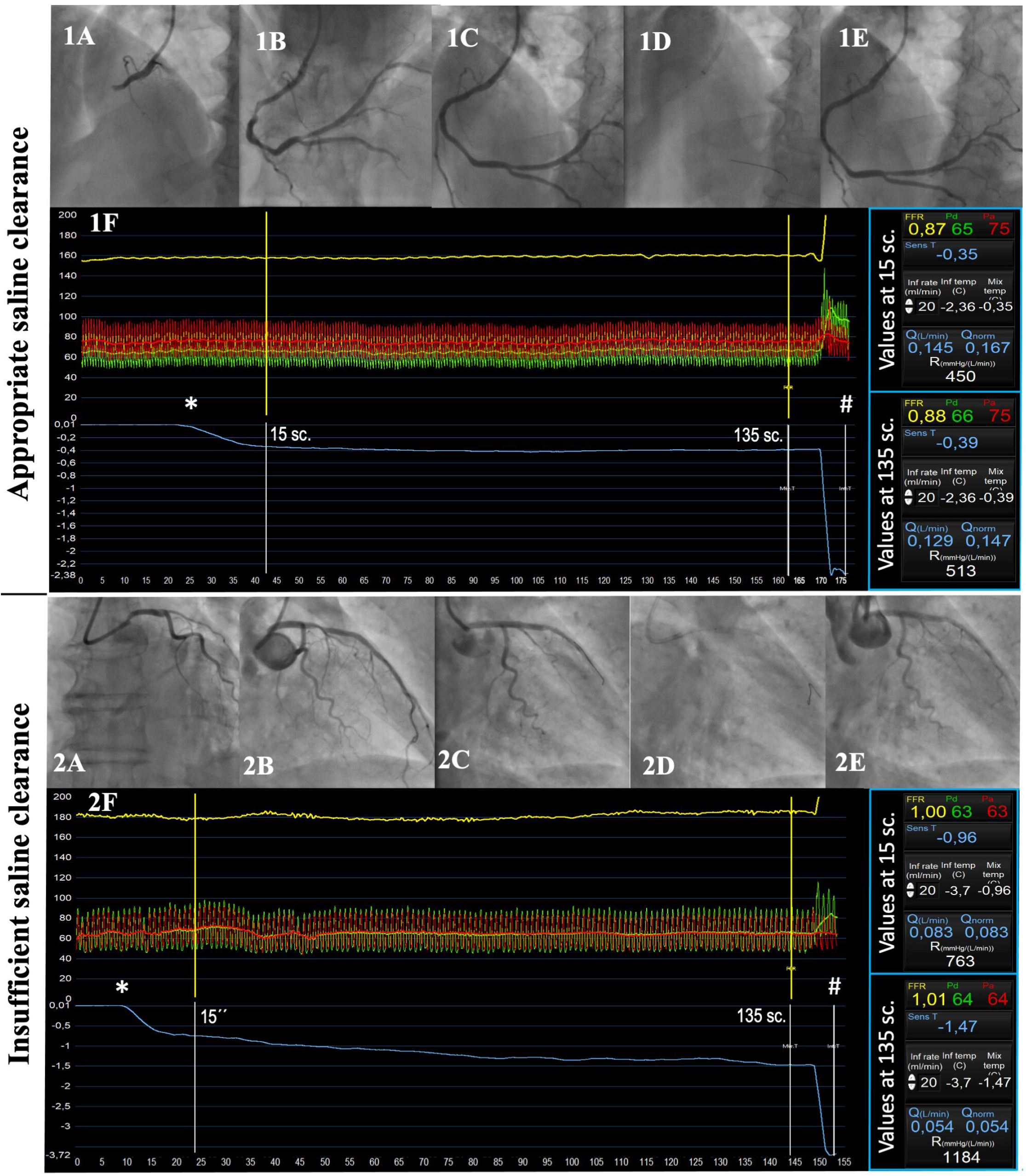
Saline-induced thermodilution patterns in patients with slow-flow. *= start of saline infusion; #= saline temperature. **Appropriate saline clearance is shown in images 1**. Acute RCA occlusion (1A). Restoration of TIMI 2 flow after thrombus aspiration (1B) with no improvement after stent implantation (1C). Flow-mediated hyperemia (1D). The thermodilution-based ACBF and MMR showed similar values at 15 seconds (145 ml/min and 450 Wood units) and at 135 seconds (129 ml/min and 513 Wood units) without variation of the distal temperature (from -0.35 to -0.39º). However, normal TIMI 3 flow restoration was observed after flow-mediated hyperemia (1E). **Deficient saline clearance is shown in figure 2** Acute LAD artery occlusion (2A). Restoration of TIMI 3 flow after thrombus aspiration (2B) followed by no re-flow (TIMI flow 1) after stent implantation (2C). Flow-mediated hyperemia (2D). The thermodilution-based ACBF and MMR showed significant changes from 15 seconds (83 ml/min and 763 Wood units) to 135 seconds (54 ml/min and 1184 Wood units) due to progressive decrease of distal temperature (from -0.96 to -1.47º) during saline infusion (2F). TIMI 2 flow was observed post-treatment (2E).

The insufficient saline clearance pattern was observed in 7 patients (18.9%) of the flow-mediated hyperemia group. Those patients presented with more severe angiographic no reflow and with lower ACBF and higher MMR before no reflow treatment than patients with appropriate saline clearance. Moreover, insufficient clearance pattern was associated with a poor response to flow-mediated hyperemia. **Table 1 of the supplemental appendix** shows the main clinical and angiographic characteristics of patients treated with flow-mediated hyperemia presenting with different thermodilution patterns.

### In-hospital clinical outcomes

No reflow was associated with a remarkable rate of in-hospital major adverse cardiac events. A total of 7 patients (10.4%) died due to cardiogenic shock (n=3), cardiac rupture (n=2), acute ventricular septal defect (n=1) and stent thrombosis (n=1). Moreover, non-fatal heart failure was observed in 18 patients (26.9%). In-hospital left ventricle ejection fraction was 44.1±9.7% (45.2±9.3% vs. 43.7±10.1%; p=0.529). Elective revascularization of non-culprit lesions was performed in 20 patients (30.0%): 19 with PCI and 1 patient with coronary artery bypass graft. **Table 5** shows the in-hospital outcomes observed in the present study.

**Table 5.** In-hospital outcomes

|  | **All patients**<br>**(n=67)** | **Pharma-**<br>**mediated**<br>**hyperemia**<br>**group**<br>**(n=30)** | **Saline-**<br>**mediated**<br>**hyperemia**<br>**group**<br>**(n=37)** | **p** |
| --- | --- | --- | --- | --- |
| **All-cause death:** | 7 (10.4%) | 2 (6.7%) | 5 (13.5%) | 0.362 |
| Cardiac rupture | 2 (3.0%) | 0 | 2 (5.4%) |  |
| Acute ventricular septal defect | 1 (1.5%) | 0 | 1 (2.7%) |  |
| Cardiogenic shock | 3 (4.5%) | 1 (3.3%) | 2 (5.4%) |  |
| Stent thrombosis | 1 (1.5%) | 1 (3.3%)* | 0 |  |
| **Non-fatal heart failure** | 18 (26.9%) | 5 (16.7%) | 13 (35.1%) | 0.090 |
| **Hemodynamic support** |  |  |  |  |
| Inotropic drugs | 9 (13.4%) | 2 (6.9%) | 7 (18.9%) | 0.158 |
| Inotropic drugs + LVAD | 4 (6.0%) | 0 | 4 (11.1%) | 0.060 |
| **Stent thrombosis** | 2 (3.0%) | 1 (3.3%)* | 1 (2.7%) | 0.880 |
| **Revascularization of non-culprit**<br>**vessels** | 20 (30.0%) | 7 (23.3%) | 13 (35.1%) | 0.140 |
| **Other non-fatal complications:** |  |  |  |  |
| Atrial fibrillation (unknown) | 2 (3.0%) | 2 (6.7%) | 5 (13.5%) | 0.196 |
| Need permanent pacemaker | 1 (1.5%) | 0 | 1 (2.7%) | 0.364 |
| Major bleeding | 3 (4.5%) | 2 (6.7%) | 1 (2.7%) | 0.435 |
| Intraventricular thrombus | 3 (4.5%) | 1 (3.3%) | 2 (5.4%) | 0.683 |

LVAD= left ventricle assist device; VSD= ventricular septal defect
\* One patient with stent thrombosis presented with in-hospital cardiogenic shock and death.

### Follow-up physiology assessment

A total of 14 patients undergoing percutaneous revascularization for non-culprit lesions (73.7%) were re-investigated with intracoronary physiology of the culprit lesion (at mean of 3.4 days after PPCI). ACBF increased from 102.8±43.7 to 142.4±57.0 ml/min (p=0.071) and MMR decreased from 926.4±420.1 to 609.1±282.2 Wood units (p=0.009). **Figure 5** shows the changes in ACBF and MMR between baseline post-treatment and follow-up.

**Figure 5.**
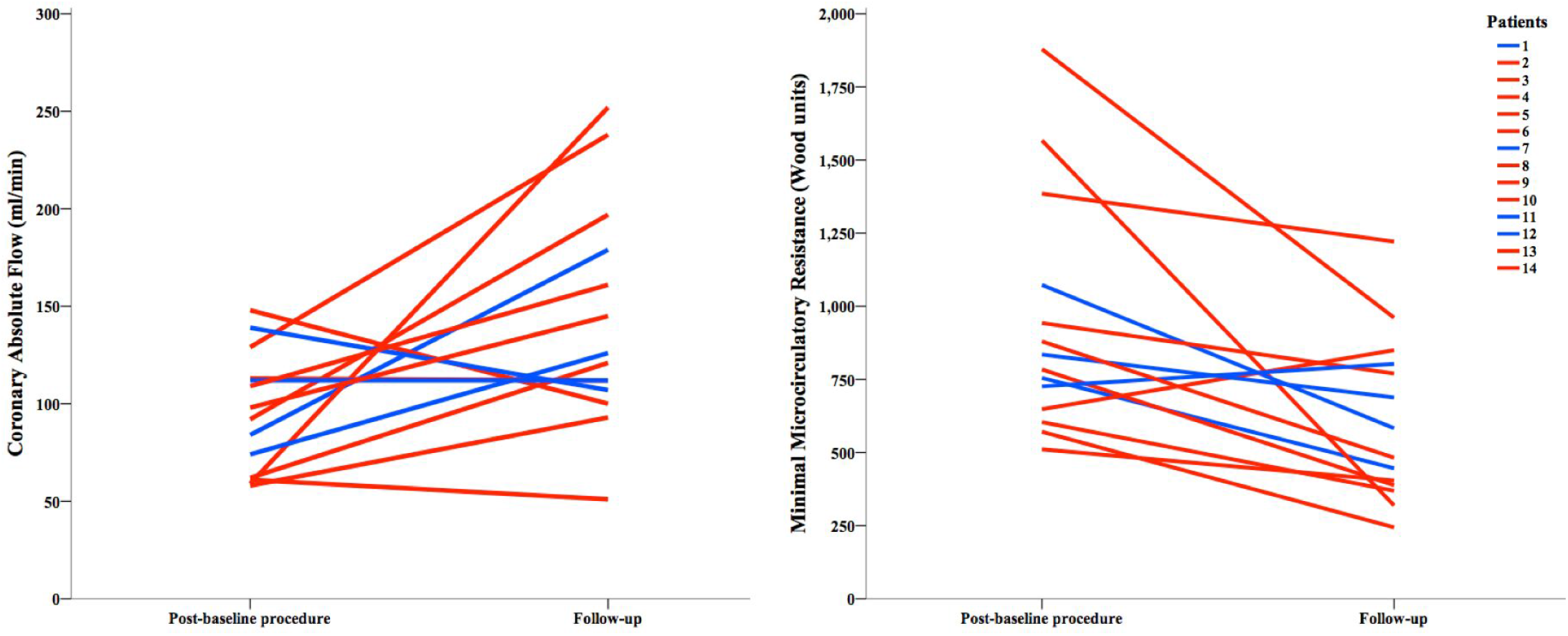
Absolute coronary blood flow and minimal microcirculatory resistance changes between baseline and follow-up procedures. Fourteen patients underwent to thermodilution-based physiologic assessment at baseline (post-intervention) and at follow-up. Baseline values were estimated at 15 seconds in the pharmacologic (blue) and at 135 seconds in the flow-mediated hyperemia group (red).

## Discussion

The main findings of this study are: 1) flow-mediated hyperemia with saline infusion via dedicated microcatheter showed similar efficacy as conventional hyperemic agents to improve the coronary flow in STEMI patients presenting with no reflow after PPCI; 2) few patients achieved normal coronary TIMI 3 flow after treatment with either pharmacologic or flow-mediated hyperemia; 3) no reflow after PPCI was associated with a high rate of in-hospital major adverse cardiac events, irrespective of the given treatment; 4) although the angiographic cTFC improved between pre and post-saline infusion in patients treated with flow-mediated hyperemia, the observed MMR values worsened during saline infusion in this group. This disagreement between angiographic and physiologic results (at the end of flow-mediated hyperemia) was probably explained by unreliable thermodilution-based physiologic values as result of insufficient saline clearance; 5) however, the thermodilution pattern observed during saline infusion was able to characterize patients with no response to flow-mediated hyperemia.

No reflow after PPCI is associated with adverse clinical outcomes ^6^. The no reflow phenomenon is mainly caused by micro-vascular obstruction due to several simultaneous mechanisms. First, coronary reperfusion of the culprit lesion is associated with micro-embolisms of thrombi and plaque detritus migrating to the microcirculatory system. Second, the microcirculation of the culprit vessel is externally compressed by inflammation of the infarcted myocardium and by micro-hemorrhages of the surrounding perivascular tissue. Finally, arteriolar spasm of the culprit vessel is often observed after reperfusion due to severe endothelial dysfunction ^8^.

Several pharmacological and mechanical interventions have been investigated to prevent and treat the slow flow phenomenon in STEMI patients (i.e., adenosine, NTP, epinephrine, hypothermia, coronary post-conditioning, remote ischemic conditioning, or tools to reduce the embolization of thrombotic material) ^5^. However, none of those therapies have been shown effective to reduce the infarcted area in large clinical trials ^7^.

The present study adds a new strategy to treat the slow flow phenomenon after PPCI. According to previous studies, saline infusion at 20 ml/min via dedicated microcatheter causes local hemolysis in the selected artery ^14^. The local release of vasodilatory agents by the disruption of red blood cells (such as adenosine and nitric oxide) is the most plausible cause of flow-mediated hyperemia ^14^. Flow-mediated hyperemia may be of interest in patients with certain hemodynamic conditions (such as severe hypotension or cardiac rhythm disorders) hampering the use of hyperemic drugs. Moreover, flow-mediated hyperemia, with continuous assessment of thermodilution-based physiologic parameters, allows to characterize the treatment response simultaneously. Patients with appropriate saline clearance pattern had better pre- and post-treatment cTFC and thermodilution-based physiologic parameters than patients with insufficient clearance pattern. Moreover, those patients also presented with better response to hyperemia than patients with insufficient saline clearance pattern (i.e., 60% of patients with appropriate clearance improved at least one degree the TIMI flow between pre- and post-treatment compared with only 29% of patients with insufficient clearance) (**Table 1 of the Supplementary appendix**). Other physiologic indices, such as the Index of Microcirculatory Resistance (IMR), have been demonstrated to identify patients with worse prognosis after PPCI ^15^. However, assessment of the IMR requires intravenous perfusion of adenosine (or analogues) and therefore, does not allow to compare different hyperemic strategies for no reflow treatment. Finally, patients undergoing new coronary angiography (i.e., in cases with scheduled treatment of non-culprit lesions), re-investigated with thermodilution-based techniques, the use of physiology was useful to investigate the coronary flow restoration few days after the index procedure. Therefore, this technique may be of interest to future research on slow flow dedicated therapies.

The present study has several limitations. First, the study failed to achieve the pre-specified sample size due to slow recruitment. Therefore, all comparisons between the study groups should be interpreted with caution and are merely hypothesis-generating. Second, the present study was designed to assess the immediate efficacy of flow-mediated hyperemia to treat the no reflow phenomenon after PPCI. A total of 40% of patients in the flow-mediated hyperemia group were treated with hyperemic drugs (as bail-out) after the study intervention because of persistent slow flow. Although the treatment cross-over did not affect the results of the co-primary endpoints (that were assessed before the cross-over), all comparisons performed after the baseline procedure (i.e., ECG ST-segment resolution or clinical outcomes) may have been influenced. Finally, as per protocol, both groups underwent to saline infusion (the pharmacologic for 15 seconds and the flow-mediated hyperemia group for 135 seconds) to assess the MMR. At 15 seconds, saline infusion has been shown to cause hyperemia in patients with chronic coronary syndrome. Therefore, the pharmacologic-mediated hyperemia group has been exposed to 2 different (pharmacologic and flow-mediated) hyperemic stimuli when the MMR was assessed at post-intervention; and this may influence the MMR results of this group.

In conclusion, the present study has not achieved the planned sample size and therefore, all results are merely hypothesis generating. According to the results of the study, patients with slow flow after PPCI treated with flow-mediated hyperemia seemed to have similar immediate angiographic and intracoronary physiologic results as patients treated with standard of care hyperemic drugs for slow flow treatment (such as adenosine and nitroprusside). Few patients achieved restoration of normal coronary flow and a remarkable number of patients presented with in-hospital major adverse cardiac events, irrespectively of the given treatment. It is noteworthy that flow-mediated hyperemia with thermodilution pattern assessment allowed the simultaneous characterization of the no reflow degree and response to hyperemia. However, the physiologic mechanisms, prevention and treatment of coronary slow flow warrants further investigations.

## Data Availability

All authors have access to the data of the study.

## Non-standard Abbreviations and Acronyms

ACBF: absolute coronary blood flow
cTFC: corrected TIMI frame count
MMR: minimal microcirculatory resistance
PPCI: Primary percutaneous coronary intervention
STEMI: ST-segment elevation myocardial infarction
TIMI: Thrombolysis In Myocardial Infarction

## References

1. Szummer K, Wallentin L, Lindhagen L, Alfredsson J, Erlinge D, Held C, James S, Kellerth T, Lindahl B, Ravn-Fischer A, Rydberg E, Yndigegn T and Jernberg T. Improved outcomes in patients with ST-elevation myocardial infarction during the last 20 years are related to implementation of evidence-based treatments: experiences from the SWEDEHEART registry 1995-2014. Eur Heart J. 2017;38:3056–3065.

2. Cequier A, Ariza-Sole A, Elola FJ, Fernandez-Perez C, Bernal JL, Segura JV, Iniguez A and Bertomeu V. Impact on Mortality of Different Network Systems in the Treatment of ST-segment Elevation Acute Myocardial Infarction. The Spanish Experience. Rev Esp Cardiol (Engl Ed). 2017;70:155–161.

3. Pedersen F, Butrymovich V, Kelbaek H, Wachtell K, Helqvist S, Kastrup J, Holmvang L, Clemmensen P, Engstrom T, Grande P, Saunamaki K and Jorgensen E. Short-and long-term cause of death in patients treated with primary PCI for STEMI. J Am Coll Cardiol. 2014;64:2101–8.

4. Yamashita Y, Shiomi H, Morimoto T, Yaku H, Furukawa Y, Nakagawa Y, Ando K, Kadota K, Abe M, Nagao K, Shizuta S, Ono K, Kimura T and Investigators CR-KAR. Cardiac and Noncardiac Causes of Long-Term Mortality in ST-Segment-Elevation Acute Myocardial Infarction Patients Who Underwent Primary Percutaneous Coronary Intervention. Circ Cardiovasc Qual Outcomes. 2017;10.

5. Rezkalla SH, Stankowski RV, Hanna J and Kloner RA. Management of No-Reflow Phenomenon in the Catheterization Laboratory. JACC Cardiovasc Interv. 2017;10:215–223.

6. Ndrepepa G, Tiroch K, Fusaro M, Keta D, Seyfarth M, Byrne RA, Pache J, Alger P, Mehilli J, Schomig A and Kastrati A. 5-year prognostic value of no-reflow phenomenon after percutaneous coronary intervention in patients with acute myocardial infarction. J Am Coll Cardiol. 2010;55:2383–9.

7. Ibanez B, Heusch G, Ovize M and Van de Werf F. Evolving therapies for myocardial ischemia/reperfusion injury. J Am Coll Cardiol. 2015;65:1454–71.

8. Ibanez B, James S, Agewall S, Antunes MJ, Bucciarelli-Ducci C, Bueno H, Caforio ALP, Crea F, Goudevenos JA, Halvorsen S, Hindricks G, Kastrati A, Lenzen MJ, Prescott E, Roffi M, Valgimigli M, Varenhorst C, Vranckx P, Widimsky P and Group ESCSD. 2017 ESC Guidelines for the management of acute myocardial infarction in patients presenting with ST-segment elevation: The Task Force for the management of acute myocardial infarction in patients presenting with ST-segment elevation of the European Society of Cardiology (ESC). Eur Heart J. 2018;39:119–177.

9. Lawton JS, Tamis-Holland JE, Bangalore S, Bates ER, Beckie TM, Bischoff JM, Bittl JA, Cohen MG, DiMaio JM, Don CW, Fremes SE, Gaudino MF, Goldberger ZD, Grant MC, Jaswal JB, Kurlansky PA, Mehran R, Metkus TS, Jr., Nnacheta LC, Rao SV, Sellke FW, Sharma G, Yong CM and Zwischenberger BA. 2021 ACC/AHA/SCAI Guideline for Coronary Artery Revascularization: Executive Summary: A Report of the American College of Cardiology/American Heart Association Joint Committee on Clinical Practice Guidelines. Circulation. 2022;145:e4–e17.

10. De Bruyne B, Adjedj J, Xaplanteris P, Ferrara A, Mo Y, Penicka M, Flore V, Pellicano M, Toth G, Barbato E, Duncker DJ and Pijls NH. Saline-Induced Coronary Hyperemia: Mechanisms and Effects on Left Ventricular Function. Circ Cardiovasc Interv. 2017;10.

11. Gomez-Lara J, Brugaletta S, Ortega-Paz L, Vandeloo B, Moscarella E, Salas M, Romaguera R, Roura G, Ferreiro JL, Teruel L, Gracida M, Windecker S, Serruys PW, Gomez-Hospital JA, Sabate M and Cequier A. Long-Term Coronary Functional Assessment of the Infarct-Related Artery Treated With Everolimus-Eluting Bioresorbable Scaffolds or Everolimus-Eluting Metallic Stents: Insights of the TROFIII Trial. JACC Cardiovasc Interv. 2018;11:1559–1571.

12. Gutierrez-Barrios A, Izaga-Torralba E, Rivero Crespo F, Gheorghe L, Canadas-Pruano D, Gomez-Lara J, Silva E, Noval-Morillas I, Zayas Rueda R, Calle-Perez G, Vazquez-Garcia R and Alfonso F. Continuous Thermodilution Method to Assess Coronary Flow Reserve. Am J Cardiol. 2021;141:31–37.

13. Rivero F, Gutierrez-Barrios A, Gomez-Lara J, Fuentes-Ferrer M, Cuesta J, Keulards DCJ, Pardo-Sanz A, Bastante T, Izaga-Torralba E, Gomez-Hospital JA, Garcia-Guimaraes M, Pijls NHJ and Alfonso F. Coronary microvascular dysfunction assessed by continuous intracoronary thermodilution: A comparative study with index of microvascular resistance. Int J Cardiol. 2021;333:1–7.

14. Gallinoro E, Candreva A, Fernandez-Peregrina E, Bailleul E, Meeus P, Sonck J, Bermpeis K, Bertolone DT, Esposito G, Paolisso P, Heggermont W, Adjedj J, Barbato E, Collet C and De Bruyne B. Saline-induced coronary hyperemia with continuous intracoronary thermodilution is mediated by intravascular hemolysis. Atherosclerosis. 2022;352:46–52.

15. Fearon WF, Low AF, Yong AS, McGeoch R, Berry C, Shah MG, Ho MY, Kim HS, Loh JP and Oldroyd KG. Prognostic value of the Index of Microcirculatory Resistance measured after primary percutaneous coronary intervention. Circulation. 2013;127:2436–41.

